# Strain Differences in Bloodstream and Skin Infection MRSA isolated between 2019-2021 in a Single Health System

**DOI:** 10.1101/2023.12.02.23299293

**Authors:** Katrina S. Hofstetter, Natasia F. Jacko, Margot J. Shumaker, Brooke M. Talbot, Robert A. Petit, Timothy D. Read, Michael Z. David

**Affiliations:** Division of Infectious Diseases, Department of Medicine, Emory University, Atlanta, Georgia, USA; Division of Infectious Diseases, Department of Medicine, University of Pennsylvania, Philadelphia, Pennsylvania, USA

**Keywords:** *Staphylococcus aureus*, whole genome sequencing, SSTI, bacteremia, clonal complex, CC5, CC8

## Abstract

*S. aureus* is a common cause of skin and soft tissue infections (SSTIs) and has recently become the most common cause of bloodstream infections (BSIs), but whether the strains causing these two clinical syndromes overlap has not been studied adequately. USA300/500 (clonal complex [CC] 8-ST8) and USA100 (CC5-ST5) have dominated among methicillin-resistant *S. aureus* (MRSA) strains in the U.S. since the early 2000s. We compared the genomes of unselected MRSA isolates from 131 SSTIs with those from 145 BSIs at a single U.S. center in overlapping periods in 2018-2021. CC8 MRSA was more common among SSTIs and CC5 was more common among BSIs, consistent with prior literature. Based on clustering genomes with a threshold of 15 single nucleotide polymorphisms (SNPs), we identified clusters limited to SSTI patients and separate clusters exclusively comprising BSI patients. However, we additionally identified 8 outbreak clusters that included at least one SSTI and one BSI isolate. This suggests that virulent MRSA strains are transmitted from person-to-person locally in the healthcare setting and the community and that single lineages are often capable of causing both SSTIs and BSIs.

## Introduction

*Staphylococcus aureus* asymptomatically colonizes the nares of 28-40% of the human population [1], [2]. *S. aureus* is a common cause of skin and soft tissue infections (SSTIs) [3], and has recently become the most common cause of bloodstream infections (BSIs) [4]. Nasal colonization of patients has been associated with an increased risk of both BSIs [5] and SSTIs [2], [3]. Colonization provides a reservoir from which *S. aureus* can access the bloodstream after a minor skin injury, trauma, surgery, or a viral infection [3], [4].

Community-associated (CA-) MRSA infections in the U.S., epidemiologically defined as those occurring in subjects with no recent healthcare exposures, have been most often reported in younger, healthy individuals [3], [6]. Healthcare-associated (HA-) MRSA infections, in contrast, are more likely to be diagnosed in older patients with comorbidities and are often invasive infections such as BSI or pneumonia [6]. Since at least 2004, the most often isolated MRSA strains in the US have belonged to clonal complex (CC)8 and CC5 [7]. USA300, which belongs to multilocus sequence type (ST) 8 (included in CC8), was initially associated with CA-MRSA infections, but since 2005 it has increasingly been recognized also as the cause of nosocomial infections [3], [6], [8]. USA300 has become the most common MRSA strain circulating in the US [9]. USA500 is a closely related CC8 strain type that is easily distinguished from USA300 by whole genome sequencing (WGS) [10], [11]. USA100 (usually ST5, which belongs to CC5) is most often a HA-MRSA strain. However, it has also been uncommonly isolated from epidemiologically defined CA-MRSA infections and from nasal carriage in subjects with no prior healthcare exposures [12]. Thus, there are no strain types that consistently distinguish between epidemiologically defined CA-MRSA and HA-MRSA infections [13].

Reports from the last 15 years indicate that the majority of MRSA SSTIs in the U.S. have their onset in the community and are caused by USA300 [3], [7], [8]. One study of strains reported in the published literature found that in 2002-2013, 62% of SSTIs were caused by USA300 and 19% by the second most common strain type, USA100 [8]. The prevalence of USA300 among BSI isolates may be increasing [14].

To understand the relationships between SSTI and BSI isolates, we performed WGS on SSTI and BSI MRSA strains from patients at two hospitals of the University of Pennsylvania from 2018-2020. We first compared the phylogenetic structure of MRSA isolates from SSTIs and BSIs. We then assessed whether any of these genomes were closely associated with one another, which would suggest recent transmission among study subjects. Among the SSTI isolates alone, three clusters were identified. We additionally identified eight clusters containing strains from both SSTI and BSI cases, suggesting epidemiological overlap and the spread of MRSA strains between patients with HA- and CA-MRSA infections within a single geographic area.

## Methods

### Patient and isolate selection

MRSA isolates were collected by the Hospital of the University of Pennsylvania (HUP) Clinical Microbiology Laboratory (CML) from routine clinical specimens obtained from outpatient clinics, emergency departments and inpatient units at HUP or the Penn Presbyterian Medical Center (PPMC). All MRSA isolates from SSTIs were biobanked in December 2018 - February 2021. After screening of the electronic medical record (EMR) to document that a MRSA isolate was from an SSTI, each source patient was contacted and offered enrollment in SEMAPHORE (Study of the Evolution of MRSA, Antibiotics, Persistence Having the Outcome of REcurrence), which is an NIH-funded study to determine risk factors for recurrent *S. aureus* infections during a 2-year period of follow-up. Approximately 28% of eligible subjects with SSTIs were enrolled prospectively in SEMAPHORE between December 2018 and February 2021 and were included in the present study. Separately, sequential patients with a MRSA BSI diagnosed in July 2018-August 2020 were included in a retrospective study (BREGOS, the MRSA Bacteremia Retrospective Epidemiologic and Genomic Outcomes Study [15].

For both SSTI and BSI patients, collection of demographic data from the electronic medical record included age, race, sex, zip code of home residence, site of collection (emergency department, outpatient clinic, or inpatient), infection type, anatomic source of infection, and epidemiological classification (CA-, HA- or community-onset, healthcare-associated [HACO]). Infections in subjects who had a first positive culture obtained >48 hours after admission to a hospital were classified as HA-MRSA. If an infection occurred in a subject <48 hours after admission to a hospital or from an outpatient, but who had one or more specific healthcare exposures (hemodialysis, surgery, nursing home stay, hospitalization within the last year or presence of an indwelling central venous catheter at the time of diagnosis), subjects were classified as having a HACO-MRSA infection. Infections were classified as CA-MRSA if the patient had none of these healthcare exposures and was cultured as an outpatient or <48 hours after hospital admission. For BSI patients, the Pitt Bacteremia Score and in-hospital mortality were recorded. The study was approved by the University of Pennsylvania IRB.

### DNA sequencing

Isolates from HUP CML were stored prospectively at -80C. To prepare DNA for sequencing, frozen cultures were streaked on blood agar plates and incubated overnight at 37°C. Single isolates were passaged onto fresh blood agar plates before single colonies were isolated for sequencing. Isolates were sequenced at the Children’s Hospital of Philadelphia Penn/CHOP Microbiome Center. Sequence libraries were prepared using the Illumina Nextera kit and sequenced by Illumina Hi-seq. Sequences for the SSTI and BSI are available on NCBI Sequence Read Archive (PRJNA918392, PRJNA751847, respectively).

### Bioinformatic analysis

The Bactopia pipeline was used to assemble fastq files, call MLST and SCC*mec* type, along with identifying variants [16]. CCs were assigned based on ST type and PubMLST. If a CC was not available for an ST the ST was used in place of the CC. All CC8 strains were classified as USA300 or USA500 based on the primer sequencing and the presence of PVL genes and arginine catabolic mobile element (ACME) [10]. To generate Maximum likelihood trees, core alignments were made using Parsnp (v 1.7.4) [17] and trees were created using IQ-Tree (v 2.2.0.3) [18]. The GGTREE package was used for visualization and figure generation [19]. Trees were rooted to GCF_000144955.1_ASM14495v1, and both a USA300 reference (GCF_022226995.1_ASM2222699v1) and USA500 reference (GCF_016916765.1_ASM1691676v1) were included. To identify outbreak clusters, we identified core genomes with fewer than 15 SNP differences, pairwise distances were calculated using Disty McMatrixface (https://github.com/c2-d2/disty). We used SCOARY2 (https://pubmed.ncbi.nlm.nih.gov/27887642/) with default parameters to assess for an association of any *S. aureus* accessory gene with BSI or SSTI.

### Statistical Analysis

Patient and isolate characteristics were compared for SSTI and BSI cohorts. The Chi-square test was used when testing for associations of demographic and clinical data with strain types with categorical data, unless sample sizes were small (<5 in a cell), in which case the Fisher exact test was performed. Comparisons of continuous data were made using the Wilcoxon rank-sum test. All statistical analysis was performed using R v4.2.2 [20].

## Results

During the study period, 131 subjects with SSTIs were enrolled, and 145 sequential subjects with a BSI were included in the BREGOS study [15]. SSTI and BSI patients did not differ significantly in distribution by age or race (**Table 1, Supplemental Figure 1**). However, SSTI patients were more likely to have CA-MRSA infections and less likely to have HA- or HACO-MRSA infections than BSI patients. SSTI patients were more likely to have been diagnosed with the infection as an outpatient (46% vs. 2%) or in the emergency department and sent home (27% vs. 3%) than BSI patients. BSI patients were nearly all diagnosed in the inpatient setting (95%) (**Table 1**). Abscesses were the most common type of SSTI (52%, n = 68), followed by surgical site infections (14%, n = 18) and infected wounds (12%, n = 16). Among BSI with a known source of infection, the most common sources were a skin site (19%) or a central venous catheter infection (14%) (**Table 2, Figure 1**). In-hospital mortality for BSI patients was 15% (22/145).

**Table 1.** Demographic and clinical characteristics of subjects with MRSA infections and clonal complexes of the causative MRSA isolates, comparing bacteremia (BSI) cases to skin and soft tissue infections (SSTIs).

| Category | SSTI n=131 (%) | BSI n=145 (%) | p-value |
| --- | --- | --- | --- |
| **Race** |  |  |  |
|  |  |  | p > 0.05 |
| White | 81 (62) | 78 (54) |  |
| Black | 43 (33) | 58 (40) |  |
| Asian | 2 (2) | 2 (1) |  |
| Other | 5 (4) | 7 (5) |  |
| **Gender** |  |  |  |
|  |  |  | p > 0.05 |
| Female | 62 (47) | 70 (48) |  |
| Male | 69 (53) | 75 (52) |  |
| **Epidemiologic Type** |  |  |  |
|  |  |  | p < 0.001 |
| HA | 5 (5) | 34 (23) |  |
| CA | 64 (49) | 17 (12) |  |
| HACO | 62 (47) | 94 (65) |  |
| **Site of Diagnosis** |  |  |  |
|  |  |  | p < 0.001 |
| Emergency Department patient | 35 (27) | 4 (3) |  |
| Inpatient | 36 (27) | 138 (95) |  |
| Outpatient | 60 (46) | 3 (2) |  |
| **Age (years)** |  |  |  |
|  |  |  | p > 0.05 |
| <50 | 52 (40) | 57 (40) |  |
| >50 | 79 (60) | 86 (60) |  |
| **Clonal complex or ST** |  |  |  |
|  |  |  | p < 0.001 |
| CC8 | 102 (78) | 79 (55) |  |
| CC5 | 20 (15) | 55 (39) |  |
| CC1 | 1 (<1) | 0 |  |
| CC6 | 1 (<1) | 0 |  |
| CC22 | 2 (1) | 0 |  |
| CC30 | 1 (<1) | 2 (1) |  |
| CC45 | 1 (<1) | 0 |  |
| CC59 | 1 (<1) | 0 |  |
| CC71 | 1 (<1) | 0 |  |
| CC72 | 0 | 6 (4) |  |
| CC78 | 0 | 2 (1) |  |
| ST87 | 0 | 1 (<1) |  |

**Table 2.** Types of skin and soft tissue infections (SSTIs) and sources of bacteremia (BSI) in this study.

| **Type of SSTI n=131 (%)** |  |
| --- | --- |
| Abscess | 68 (52%) |
| Surgical Site Infection | 18 (14%) |
| Infected Wounds | 16 (12%) |
| Cellulitis | 15 (11%) |
| Folliculitis | 3 (2%) |
| Impetigo | 3 (2%) |
| Pustule | 3 (2%) |
| Otitis Externa | 1 (<1%) |
| Paronychia | 1 (<1%) |
| Cyst | 1 (<1%) |
| Pyomyositis | 1 (<1%) |
| Tenosynovitis | 1 (<1%) |
| **Type of BSI n=145 (%)** |  |
| Skin site | 27 (19%) |
| Central venous catheter infection | 20 (14%) |
| Surgical site | 7 (5%) |
| Device infection | 5 (3%) |
| Arteriovenous Graft | 4 (3%) |
| Respiratory source | 3 (2%) |
| Urinary source | 3 (2%) |
| Other | 5 (3%) |
| Unknown | 71 (49%) |

**Figure 1.**
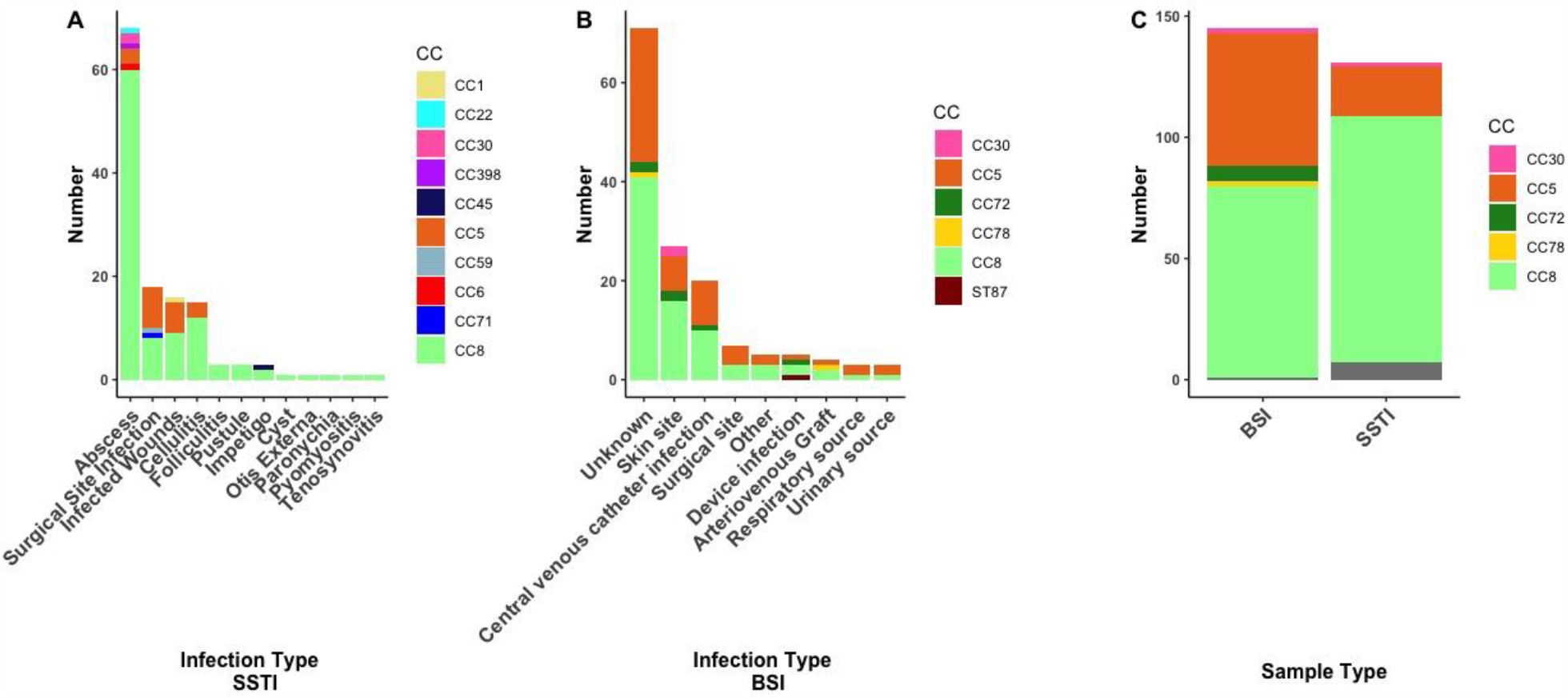
A) Distribution of infection types among the skin and soft tissue infections (SSTIs), stratified by clonal complex. B) Distribution of infection types among the bloodstream infections (BSIs), stratified by clonal complex. C) Clonal complex distribution comparing SSTI and BSI strains.

### Comparing strain types among SSTI and BSI MRSA Isolates

SSTI isolates were assigned to 14 STs by genome-based MLST. CC8 and CC5 were the most abundant clonal complexes among the SSTI isolates, accounting for the vast majority of all MRSA strains, with CC8 being the most prevalent (102/131, 78%) and CC5 second most (20/131, 15%) (**Table 1**). Among abscesses, CC8 strains were by far the most common (60/68, 88%, p-value >0.01) (**Figure 1**). BSI isolates belonged to 5 CCs (CC8, CC5, CC72, CC78, and CC30), and 1 ST was not included in a CC (ST87). The BSI isolates were in 13 STs. As for SSTI isolates, the majority of BSI isolates were CC5 or CC8 (55/145, 38%, and 79/145, 54%, respectively), although the CC8 strains made up a significantly larger proportion of the SSTI than the BSI strains (p <0.1) (**Table 1, Figure 1**).

We compared the genomes of the 131 SSTI isolates to 145 BSI isolates [15]. All of the CC8 strains were subtyped into either USA300 (n=101 SSTI/n= 69 BSI) or USA500 [10] (n=1 SSTI/n= 10 BSI). SSTI and BSI strains belonging to the same ST did not form separate subclades but instead were polyphyletic (**Figure 2**). The principal difference between the groups of strains was in the balance of the genotypes: CC8 were overrepresented in SSTI (102/131, 78%, p value >0.01) compared to CC5 (21/131, 16%) (**Figure 1, Table 1**). CC5, in contrast, were more often found as a cause of BSIs than of SSTIs (55/145, 38%).

**Figure 2.**
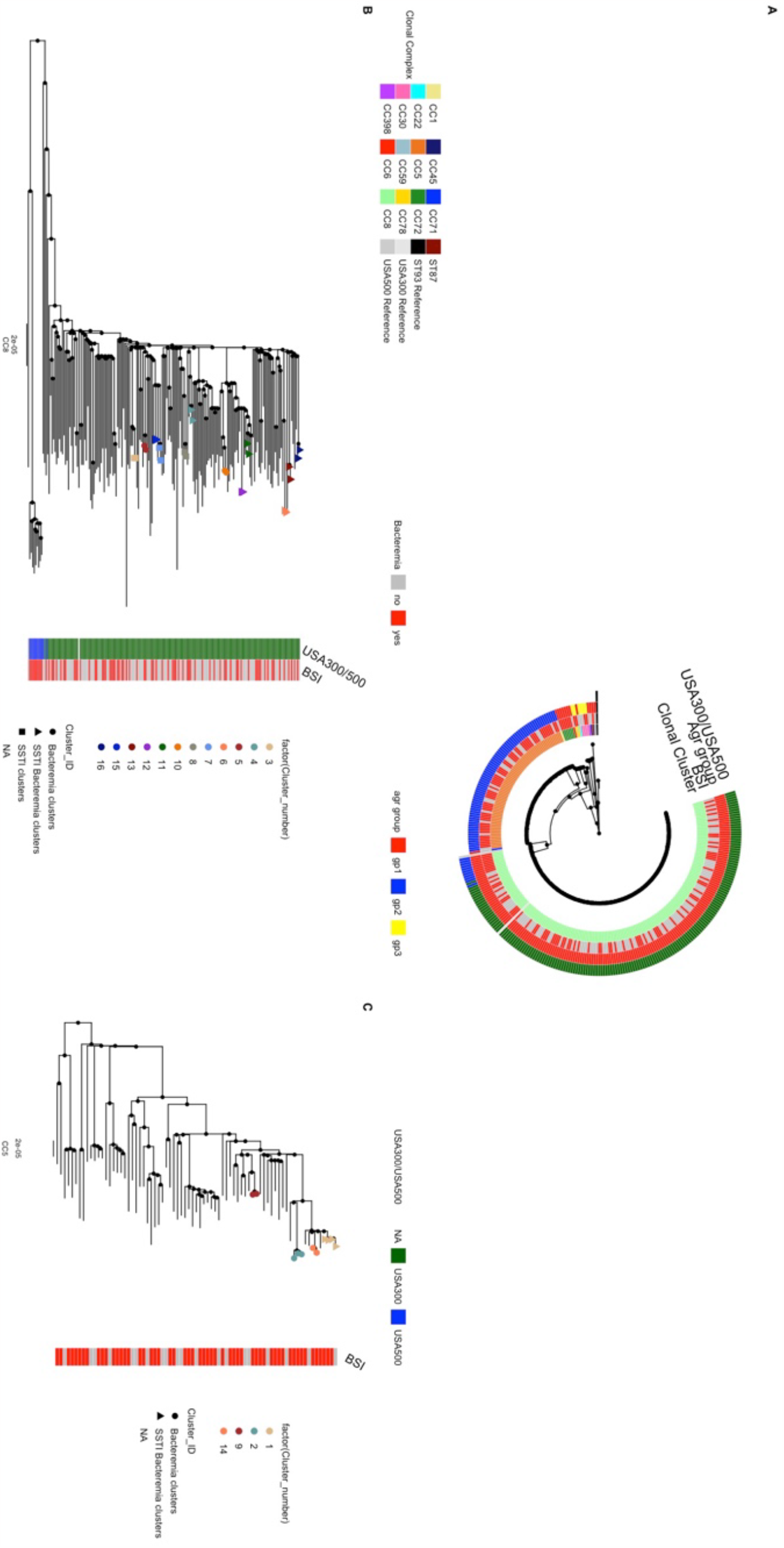
A) Maximum likelihood tree of all sequenced samples. Heatmap depicts clonal complexes, if the samples are from a bloodstream infection (BSI), and agr group. B) CC8 cluster only, C) CC5 cluster only, branch tips indicate outbreak clusters > 15 SNPs. Clusters with only BSI strains are represented with a circle, clusters with only strains from skin and soft tissue infections (SSTIs) are represented with a square, and clusters containing both BSI and SSTI strains are represented by triangles.

Comparing the antimicrobial susceptibilities of BSI isolates to SSTI isolates, the patterns are quite similar overall. However, BSI isolates were less likely to be susceptible to clindamycin (59% vs. 73%), erythromycin (11% vs. 17%), and trimethoprim/sulfamethoxazole (93% vs. 100%) (**Table 3)**. These associations were confounded by the different strain backgrounds associated with SSTI/BSI and there was not a significant association between AMR genes and disease state when adjusting for phylogeny using the pangenome GWAS tool SCOARY2.

**Table 3.** Susceptibility of MRSA to antimicrobial agents, comparing skin and soft tissue infection (SSTI) isolates to bacteremia (BSI) isolates.

|  | SSTI n=131 (%) |  | BSI n=145 (%) |  |
| --- | --- | --- | --- | --- |
| Drug name | Number tested | Susceptible | Number test | Susceptible |
| Ampicillin | 130 | 0 | 145 | 0 |
| Cefazolin | 130 | 0 | 145 | 0 |
| Penicillin | 131 | 0 | 145 | 0 |
| Oxacillin | 131 | 0 | 145 | 0 |
| Gentamicin | 131 | 126 (96) | 144 | 142 (99) |
| Ciprofloxacin | 131 | 38 (29) | 144 | 45 (31) |
| Levofloxacin | 131 | 38 (29) | 145 | 45 (31) |
| Moxifloxacin | 131 | 39 (30) | 144 | 46 (32) |
| Erythromycin | 131 | 22 (17) | 145 | 16 (11) |
| Clindamycin | 131 | 95 (73) | 145 | 86 (59) |
| Quinupristin-Dalfopristin | 130 | 130 (100) | 144 | 144 (100) |
| Linezolid | 130 | 130 (100) | 144 | 144 (100) |
| Vancomycin | 131 | 131 (100) | 145 | 145 (100) |
| Tetracycline | 131 | 120 (92) | 145 | 125 (86) |
| Tigecycline | 131 | 128 (98) | 145 | 139 (96) |
| Nitrofurantoin | 131 | 129 (98) | 144 | 144 (100) |
| Rifampicin | 131 | 130 (99) | 145 | 141 (97) |
| Trimethoprim/Sulfamethoxazole | 131 | 131 (100) | 145 | 135 (93) |
| Daptomycin | 0 | 0 | 130 | 130 (100) |
| Ceftaroline | 0 | 0 | 5 | 5 (100) |
| Cefoxitin | 130 | 0 (0) | 144 | 2 (1) |

### Transmission clusters among SSTI and BSI strains

While it has been reported that SSTI can serve as a reservoir for BSI, it is not known if MRSA causing BSIs and MRSA causing SSTIs, even within the same clonal complexes, are distinct from one another. We thus investigated, among isolated genomes from our study, how closely strains from different patients with BSIs and SSTIs were related. Several studies have indicated that a 15 SNP difference threshold can be used to cluster isolates into epidemiologically linked groups that shared the same common ancestor in the past few years [15], [21]–[25]. In our combined group of 276 isolates from SSTI and BSI, we identified 16 clusters containing between 2 and 4 isolates using the 15 SNP threshold, representing 13% (35/276) of all isolates. The correlation of pairwise distances between the phylogenetic tree and the distance matrix was 0.997 (**Supplemental Figure 2**). Three clusters contained only strains from SSTIs (all CC8) and in five clusters all strains were from BSIs (two CC8 and three CC5). Eight outbreak clusters contained at least one BSI isolate and one SSTI isolate, seven of which were CC8 and one was CC5 (**Figure 2 B,C, Table 4**). Comparisons of the electronic medical records revealed that one cluster that contained both a BSI and SSTI came from the same patient (cluster 15), while the remaining seven came from different patients. This result suggested frequent epidemiological overlap between patients with SSTIs in the community and BSI in hospitals.

**Table 4.**
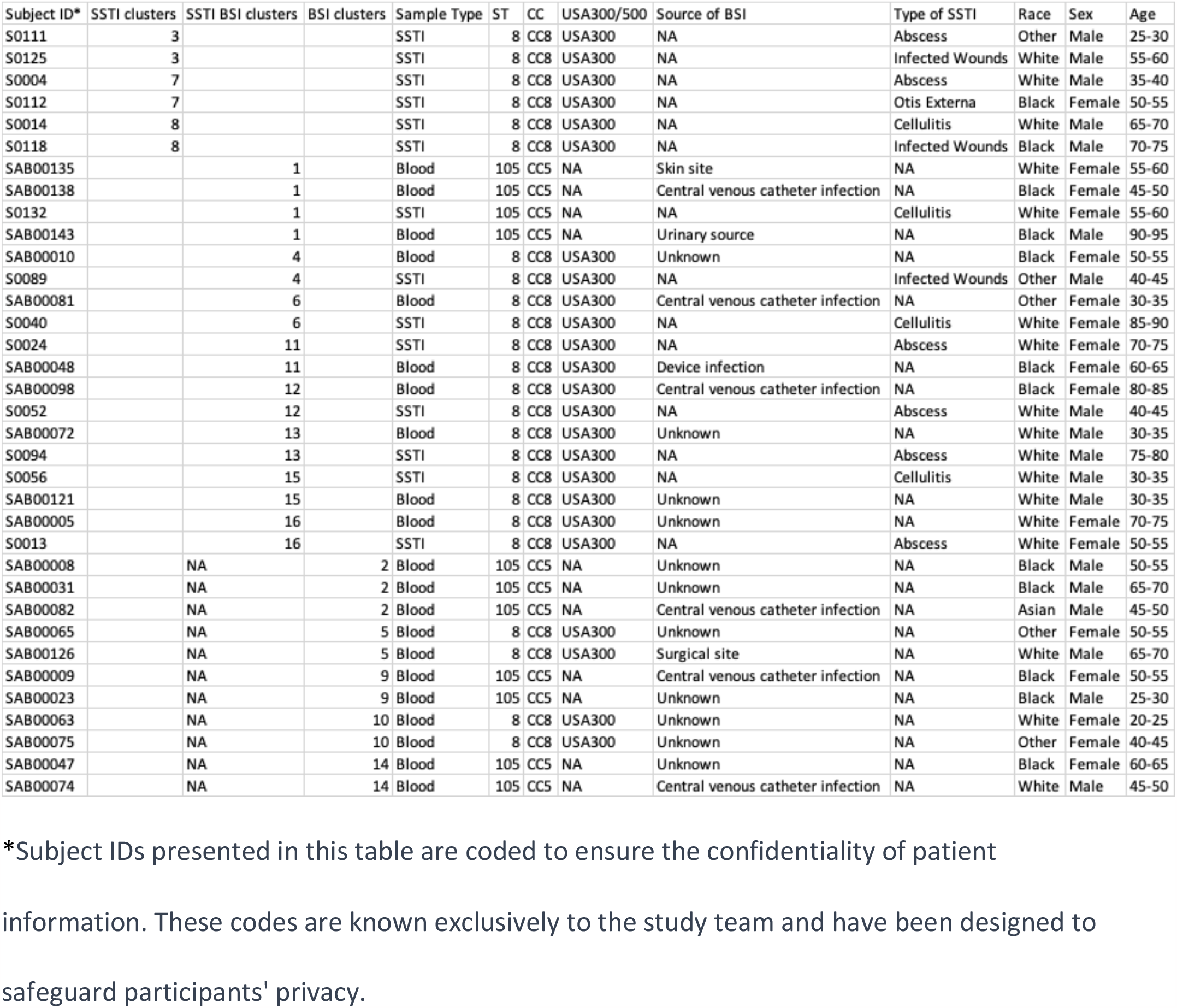
Demographic and strain data for all MRSA outbreak clusters.

## Discussion

The results of this study highlight divergent trends in infections caused by the two most common *S. aureus* clonal complexes currently isolated in the USA. CC8 isolates were strongly associated with SSTI and CC5 with BSIs. CC8 isolates that are subtyped to USA300 were the most common strain type in our study population.

The USA300 MRSA strain likely emerged in the early 1970s and spread rapidly during the 1990s and 2000s across North America to achieve its current predominant position among MRSA strain types [7], [26]. One study of 224 ST8 genomes suggested that the USA300 clone descended from an ST8 progenitor originating in central Europe in the 19th century, with an introduction to North America in the early 20th century, while gradually acquiring its typical genomic and virulence characteristics such as PVL, SCCmec type IVa, and the arginine catabolic mobile element (ACME) [27]. Another study of USA300 MRSA genomes suggested that the USA300 clone first emerged on the East Coast of the US, and perhaps in Pennsylvania [28].

Although USA300 MRSA was originally identified as a “CA-MRSA strain”, it was soon also recognized as a nosocomial pathogen as early as 2005 [6], [8], [29], [30]. Indeed, USA300 has since become the leading cause of BSI in the US, which are usually classified as HA- or HACO-MRSA infections [4]. With this shift from the community to the healthcare setting, we previously demonstrated among 616 sequential MRSA isolates from the University of Chicago Medical Center in 2004-5 that there was already a poor correlation between the typical genotypic characteristics of “CA-MRSA” strain types, such as ST8, carrying SCC*mec* type IV, PVL gene carriage, or lack of a multidrug resistance phenotype with the CDC’s epidemiologic criteria for CA-MRSA infections [13].

In the current study, carried out in Philadelphia (750 miles from Chicago), and with isolates collected about 15 years later, the ST5 (USA100) and ST8 (USA300 or USA500) strains continue to dominate among MRSA isolates obtained from both blood and SSTIs. Many other U.S. and Canadian studies with isolates collected between 2003 and 2018 have shown similar results, with a generally increasing percent of BSIs and SSTIs caused by USA300/USA500 and a decreasing percent caused by USA100 [14], [31]–[40]. In a cohort of 276 MRSA SSTI and BSI patients at a single U.S. institution in 2018-2021, CC5 and CC8 strains predominated. In fact, 93% of all SSTI strains were either CC5 or CC8, and CC8 was significantly more common among SSTI than among BSI patients. Many epidemiologic characteristics of patients and their MRSA isolates were consistent with literature from the past 15 years, indicating only slowly shifting trends in relative strain prevalence during this period. Remarkably, we found that closely clonally related strains of MRSA caused both BSIs and SSTIs in different patients in our cohort, suggesting that the pathogenesis of invasive infections in very different human tissues may not require highly adapted strains. We do not yet know to what extent genetic differences between the clonal complexes (e.g., carriage of virulence genes such as arginine catabolic mobile element or PVL) play a role in the observed differences in sites of infection or if these differences.

We found in this study that abscesses are the most common type of SSTI, and they were significantly enriched among the infections caused by CC8 strains, an observation consistent with previous literature on CA-MRSA strains [41]. Also, in patients greater than 50 years of age who presented with an SSTI, we observed a higher prevalence of CC5 strains compared with younger patients. This trend could not be explained by differences in any specific infection type, although we did find that patients greater than 50 years old in our cohort were more likely than younger patients to have surgical site infections, and infected ulcers and wounds. The older population is likely to have had more healthcare-associated risk factors than those under 50, and CC5 may be more prevalent in the older cohort because CC5 strains tend to cause infections more often in hospital, rather than community, settings.

There have been few previous studies examining the genomes of MRSA infections from different anatomic sites at a single center. We identified potential MRSA transmission clusters using a conservative threshold of 15 or fewer SNPs. Three clusters were of only SSTI strains, suggesting small-scale outbreaks among the patients. There were also (five) clusters of only BSI strains, representing probable healthcare transmission, as reported by Talbot et al [15].

Surprisingly, given that we showed BSI and SSTI were commonly caused by different *S. aureus* clones, we found that 50% of the clusters contained both disease types. This result suggested epidemiologic linkage between BSI and SSTI may be more frequent than previously realized. There is literature that discusses potential BSI reservoirs being concurrent SSTI [14] and colonization of the nares [5]. A prior SSTI within the last year also increased the risk of a BSI [14]. Thus, it is possible that some BSI patients in our study became bacteremic after a MRSA SSTI; however, only a small number of patients had a known SSTI as a source of BSI in our cohort.

Limitations of our study include the relatively small size of the cohorts examined and the lack of epidemiologic data about potential interactions among subjects. Also, the study was performed only at a single center and thus may not be generalizable.

Our work highlights the importance of considering strain background in future studies of *S. aureus* to understand the relative roles of strain and patient characteristics in determining sites of infection.

## Data Availability

All data is available on NCBI Sequence Read Archive PRJNA918392, PRJNA751847.

## Conflicts of interest

The authors report no conflicts of interest.

## Funding

The authors were supported by the National Institute of Health (NIH) (grant number RO1 AI139188)

## Notes

### Competing Interest Statement

The authors have declared no competing interest.

### Author Declarations

Ethics committee/IRB of University of Pennsylvania gave ethical approval for this work. Protocols Genomics of S. aureus Colonization after Initial and Recurrent Skin Infections and the Impact of Antibiotics (831208) and Retrospective MRSA Bacteremia (843095) were approved August 2018 and May 2020, respectively.

