## Supplementary material for "Strain Differences in Bloodstream and Skin Infection MRSA isolated between 2019-2021 in a Single Health System": Suplemental Figures

**Supplemental Figures**

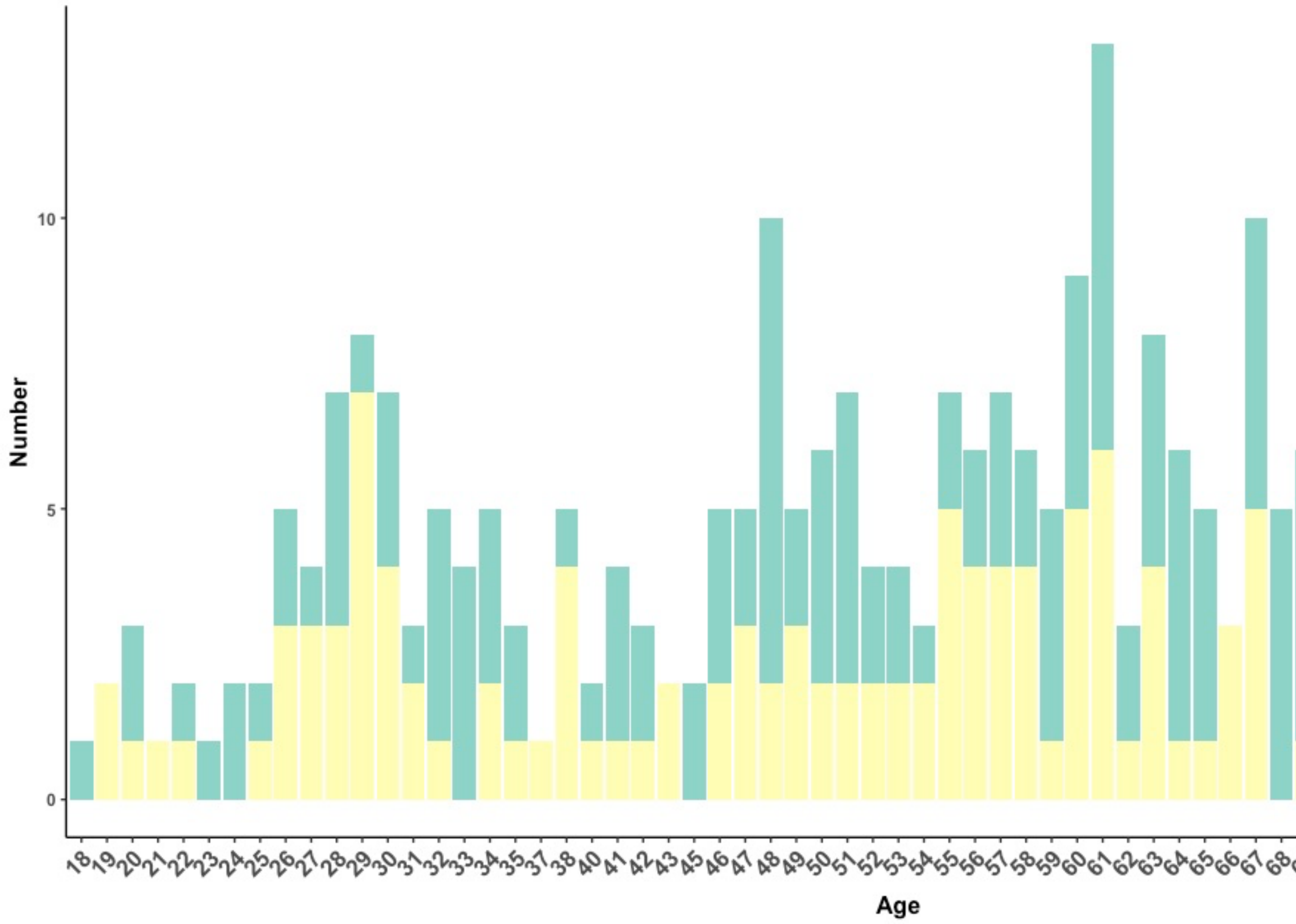


Supplemental Figure 1. Age distribution of subjects with a skin and soft tissue infection

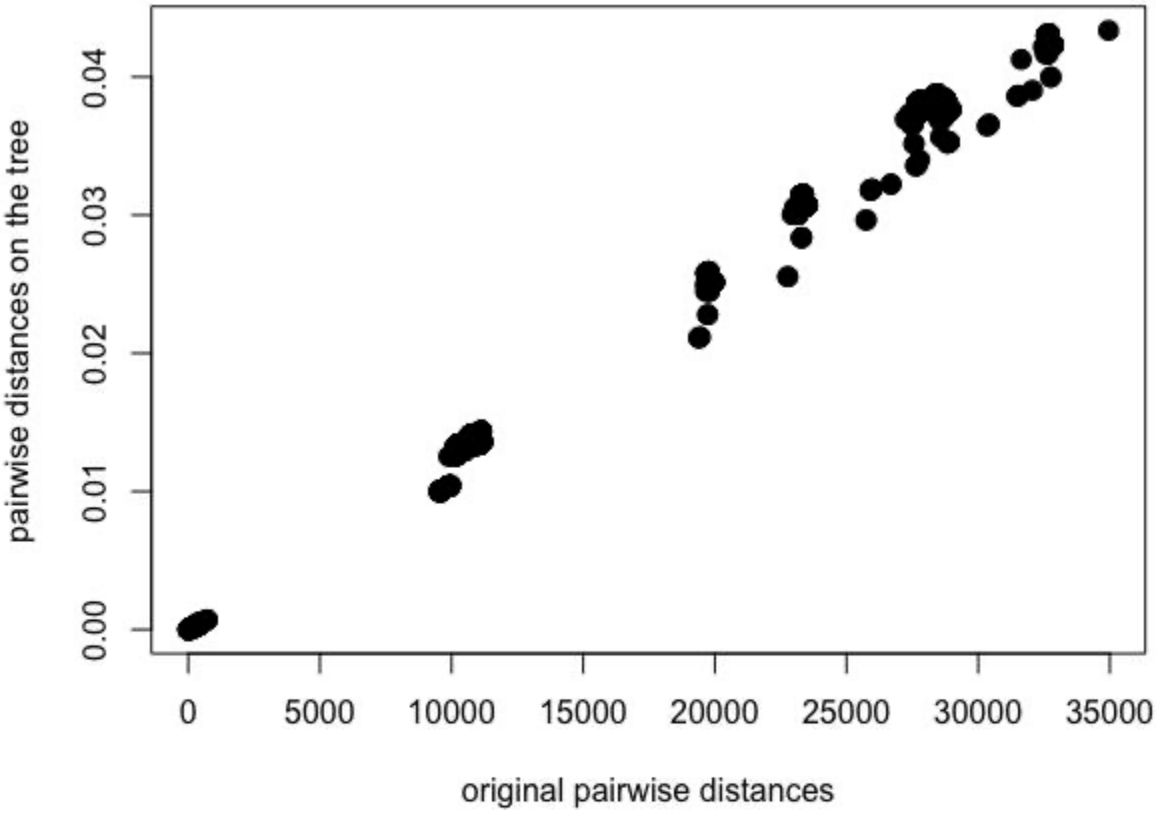


Supplemental Figure 2. Cophenetic graph showing the relation of pairwise distances on the phylogenetic tree. The correlation between pairwise distances on tree and pairwise distances from the distance matrix is 0.997.
